# Associations of functional HLA class I groups with HIV viral load in a heterogeneous cohort

**DOI:** 10.1101/2022.06.21.22276431

**Authors:** Adrian G. Zucco, Marc Bennedbæk, Christina Ekenberg, Migle Gabrielaite, Preston Leung, Mark N. Polizzotto, Virginia Kan, Daniel D. Murray, Jens D. Lundgren, Cameron R. Macpherson, the INSIGHT START study group

**Affiliations:** PERSIMUNE Center of Excellence, Rigshospitalet, Copenhagen, Denmark; Virus Research and Development Laboratory, Virus and Microbiological Special Diagnostics, Statens Serum Institut, Copenhagen, Denmark; Center for Genomic Medicine, Copenhagen University Hospital, Copenhagen, Denmark; Clinical Hub for Interventional Research, College of Health and Medicine, The Australian National University, Canberra, Australia; George Washington University, Veterans Affairs Medical Center, Washington D.C, U.S.A.

## Abstract

Human Leucocyte Antigen (HLA) class I alleles are the main host genetic factors involved in controlling HIV-1 viral load (VL). Nevertheless, HLA diversity has proven a significant challenge in association studies. We assessed how accounting for binding affinities of HLA class I alleles to HIV-1 peptides facilitate association testing of HLA with HIV-1 VL in a heterogeneous cohort from the Strategic Timing of AntiRetroviral Treatment (START) study. We imputed HLA class I alleles from host genetic data (2,546 HIV+ participants) and sampled immunopeptidomes from 2,079 host-paired viral genomes (targeted amplicon sequencing). We predicted HLA class I binding affinities to HIV-1 and unspecific peptides, grouping alleles into functional clusters through consensus clustering. These functional HLA class I clusters were used to test associations with HIV VL. We identified four clades totalling 30 HLA alleles accounting for 11.4% variability in VL. We highlight HLA-B*57:01 and B*57:03 as functionally similar but yet overrepresented in distinct ethnic groups, showing when combined a protective association with HIV+ VL (log, β −0.25; adj. p-value < 0.05). We further demonstrate only a slight power reduction when using unspecific immunopeptidomes, facilitating the use of the inferred functional HLA groups in other studies. The outlined computational approach provides a robust and efficient way to incorporate HLA function and peptide diversity, aiding clinical association studies in heterogeneous cohorts. To facilitate access to the proposed methods and results we provide an interactive application for exploring data.

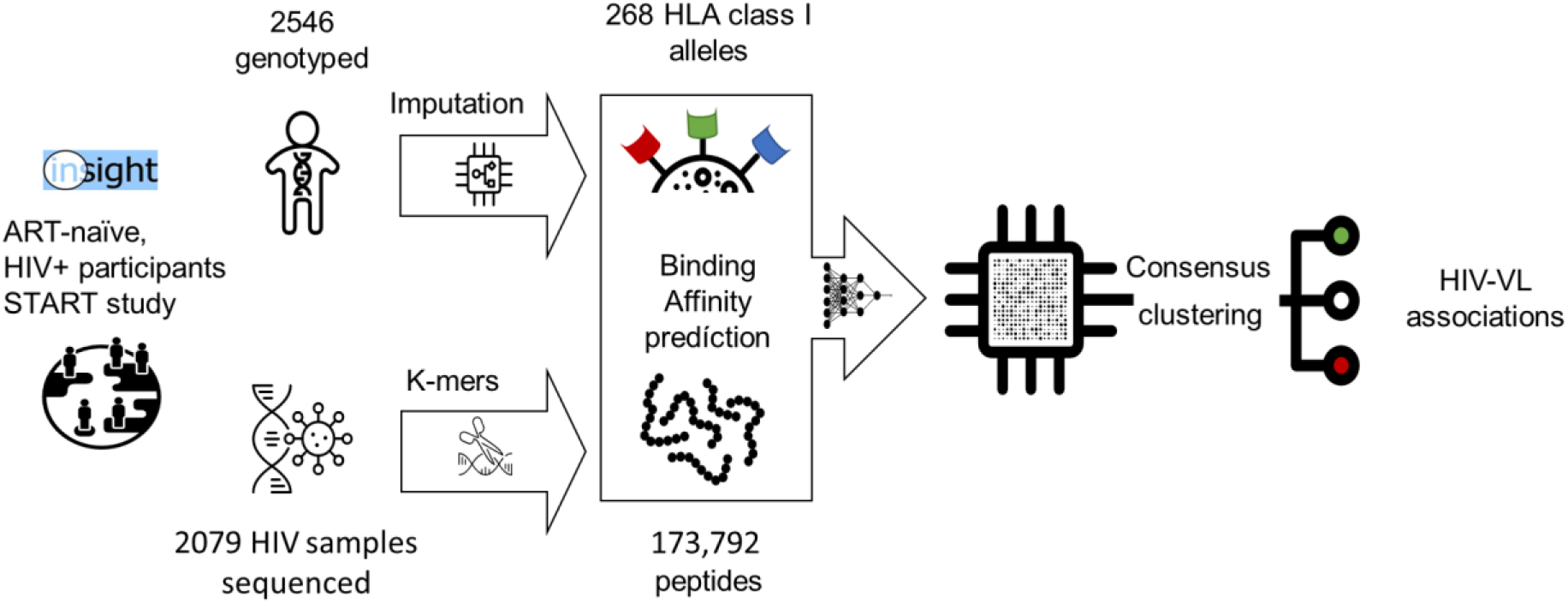

## Introduction

The Human Leucocyte Antigen (HLA) is a critical component of the host immune response. HLA Class I alleles mediate the anti-viral response through the presentation of intracellular viral peptides for recognition by Cytotoxic T cells. This mechanism is critical to the host’s defence against diverse pathogens, which is why it is among the most genetically diverse regions in the human genome as evidenced by its association with our variable response to infectious disease [1]. In the context of HIV, HLA alleles are the canonical host genetic factors associated with viral load (VL)[2], altogether contributing up to 12% of its variability [3]. Viral diversity, on the other hand, is thought to explain two to four times as much (20%-46%) [4]. The combined relevance of both host and viral genomics suggests the necessity for the simultaneous analysis of both in the context of infectious disease[5]. Different approaches have been used to study this interaction such as genome-to-genome [6] or peptidome-wide associations [7]. In addition, when considering the interplay between host HLA alleles and viral peptides, their association could be explained functionally in terms of epitope binding and presentation.

Analysis of genetic variance within the HLA region and its associations with clinical outcomes is challenged by population-dependent distributions and the diversity of HLA haplotypes. Reaching statistical power within this region is difficult and while accounting for the effects of this important immunological region is necessary, it is often ignored altogether. Yet, the function of HLA is conserved as a core component of the immune response [8], [9]. This suggests there may be shared functional features within and between populations, even in the presence of genotypic plasticity. Understanding the structure of this mutual information could prove relevant for the study of host-pathogen interactions where high mutation rates are observed such as in HIV [10].

Immunopeptidomes have been used to estimate functional similarities between HLA alleles in what was initially denominated HLA supertypes [11]. These functional groups are based on the propensity of various alleles to bind similar sets of peptides. The algorithms used to estimate these binding profiles have gradually improved over time [12], [13]. However, the consideration of HLA functional groups in the context of Genome-Wide Association Studies (GWAS) has largely been overlooked [14].

In this study, we considered the ability of HLA proteins to functionally bind peptides leveraging the cost-effectiveness of high-throughput genotyping as opposed to directly assaying HLA function. We provide a framework based on state-of-the-art computational methods for the study of predicted immunopeptidomes and functional HLA groups in the context of HIV-1 infection. In doing so, we demonstrate the increased statistical power that is gained by moving away from a purely genetic approach to a functional one with implications for studying the immune response either directly or as a confounder. We processed HIV immunopeptidomes that incorporate intra-host viral diversity and imputed HLA alleles from the same geographically diverse cohort of people living with HIV (PLWH) and antiretroviral therapy (ART) naïve participants from the Strategic Timing of Anti-Retroviral Treatment (START) trial [16]. Interactive results of this work and complementary data are available via a web application (https://persimune-health-informatics.shinyapps.io/PAW2022ZuccoHLA_HIV_INSIGHT/).

## Methods

### Ethics

Host and viral samples in this study were extracted and analyzed from participants in the START clinical trial (NCT00867048) [16], conducted by the International Network for Strategic Initiatives in Global HIV Trials (INSIGHT) and the Community Programs for Clinical Research on AIDS (CPCRA). Written consent for the study and genetic analyses were obtained from the participants and approved by participants’ site ethics review committees.

### HLA class I alleles

Imputation of HLA class I alleles (HLA-A, HLA-B and HLA-C) for 2546 genotyped ART-naïve, HIV+ participants was performed with HIBAG at 4-digit resolution [17]. Full details of the imputation process and quality control are described in previous publications [2], [18]. A multi-ethnic pre-trained model was used for imputation with a minimum out-of-bag accuracy of over 90% for all loci. HLA diversity was measured by the inverse Simpson index based on the HLA allele frequencies per locus and country. This index represents the complement of the probability that two participants would have the same HLA allele for a selected locus in a country.

### Immunopeptidomes and binding affinity prediction

Plasma samples were obtained for a subset of 2079 ART-naïve, HIV+ participants from 21 countries enrolled in the START study. Viral RNA was sequenced, paired-end, using Illumina MiSeq and covered two amplicons in the HXB2 genome positioned 1485-5058 and 5967-9517. The sample preparation, library preparation, sequencing procedure, and detailed quality controls have been described previously [19]. Raw reads were fragmented into 27-mers using KAT [20] and those with a count higher than 1 were translated into peptide sequences of 9 amino acids length to fit the mean length of HLA Class I epitopes. Peptides were mapped to 10 major HIV proteins (Asp, Gag-Pol, Nef, Vpr, Vpu, gp160, Vif, Pr55, Rev, Tat) from NCBI RefSeq NC001802.1 (Supplementary file 1) using BLAST 2.8.1 (blastp-short). Hits with an E-value > 1E-05 were excluded to remove low-quality k-mers by considering exact matches. To compare HIV peptidomes with random sequences, a set of half-million 9-mers were generated by processing the same number of random protein sequences from Uniprot. Binding affinities for 268 class I HLA alleles to both HIV and random peptidomes using NetMHCpan 4.0 [15]. Three different immunopeptidome subsets were generated by (i) selecting peptides from the top 10% binding affinities, (ii) 10% most variable peptides, and (iii) peptides that potentially bind to at least 10% of the alleles using <500nM as general threshold [12].

### Consensus clustering

Hierarchical clustering was implemented using two different linkage functions. Average linkage was used for measuring relationships between HLA alleles represented by dissimilarity defined as cosine, correlation and Euclidean distances. For clustering based on Ward linkage, cosine and correlation distances were corrected by the square root to satisfy the triangular inequality necessary to operate in Euclidean space [21]. To generate an ensemble of clustering solutions, we employed consensus clustering to mitigate bias from the subset, distance metric, or chosen linkage function [22]. A consensus matrix (C_ij_) of size (n x n) was built where each element is the number of times an i^th^ allele clustered together with a j^th^ allele [23] at a varying total number of clusters selected (3 to 160). The consensus matrix was then processed by hierarchical clustering with average linkage after transforming the values into dissimilarity scores (1 – C_ij_). Clustering was performed in Python 3.7.1 using the Scipy library [24].

### Statistical analyses

Associations of log_10_-transformed VL with each node of the consensus tree were tested by linear regression and adjusted by sex, self-reported race, and country. Tested HLA alleles had to be present in more than 10% of the participants. Multiple testing was controlled by a Benjamini-Hochberg procedure using a q-value < 0.05 to identify associations. Analyses were performed in R v3.6.0 [25].

### Data visualization and availability

Consensus clustering dendrograms and association coefficients were depicted using Interactive Tree of Life (iTOL) [26] and tanglegrams, which were generated by the *dendextend* R package [27]. We provide a flexible visualization of peptide-to-HLA binding profiles across viral proteins. Data downloads and access to supplementary materials are made available through the app (https://persimune-health-informatics.shinyapps.io/PAW2022ZuccoHLA_HIV_INSIGHT/).

## Results

### Baseline characteristics for the START cohort

We used baseline data and the genotypes of 2,546 participants from the START trial. Next-generation sequencing of HIV samples was retrieved for a subset of 2,079 participants. All participants included in the trial were asymptomatic HIV+ and ART-naïve with two CD4+ cell counts >500/μL at least 14 days apart within 60 days of enrollment in the trial. Baseline characteristics for study participants can be found in Table 1.

**Table 1.**
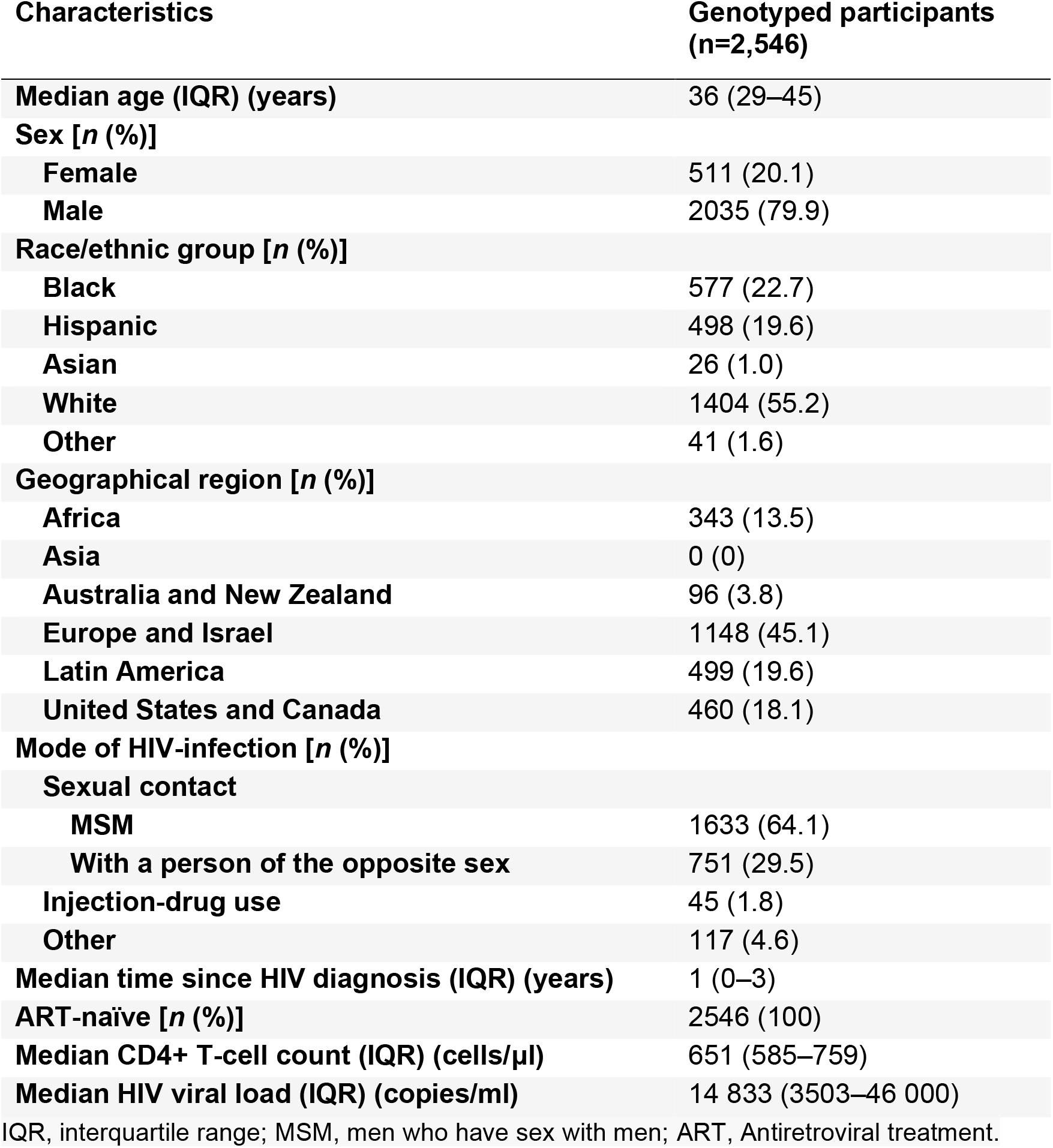
Demographic characteristics of the START cohort at study entry.

### Prediction of patient-derived HIV immunopeptidomes

Our approach for immunopeptidome generation initially yielded 9.88 × 10^7^ peptide 9-mers. After filtering and mapping with BLAST to ten major HIV proteins, a total of 173,792 9-mers were considered. This accounted for a 99.22% coverage across the reference proteome. Among the final list, we observed 136 best-defined CTL/CD8+ epitopes from Los Alamos (version 2019-11-20; Supplementary file 2).

### Data exploration

To facilitate the exploration of the results, a web application was developed (Figure 1) providing tools to navigate interactively the global and local diversity of imputed HLA alleles, HIV subtype-derived peptides, and their corresponding binding profiles. Shannon and Simpson diversity indices can be directly visualized on the world map highlighting the geographical diversity of the cohort. This interactive tool facilitates the further examination of specific HLA allele, HIV subtype, or peptide frequencies for the entire cohort and options to explore details at a country level.

**Figure 1.**
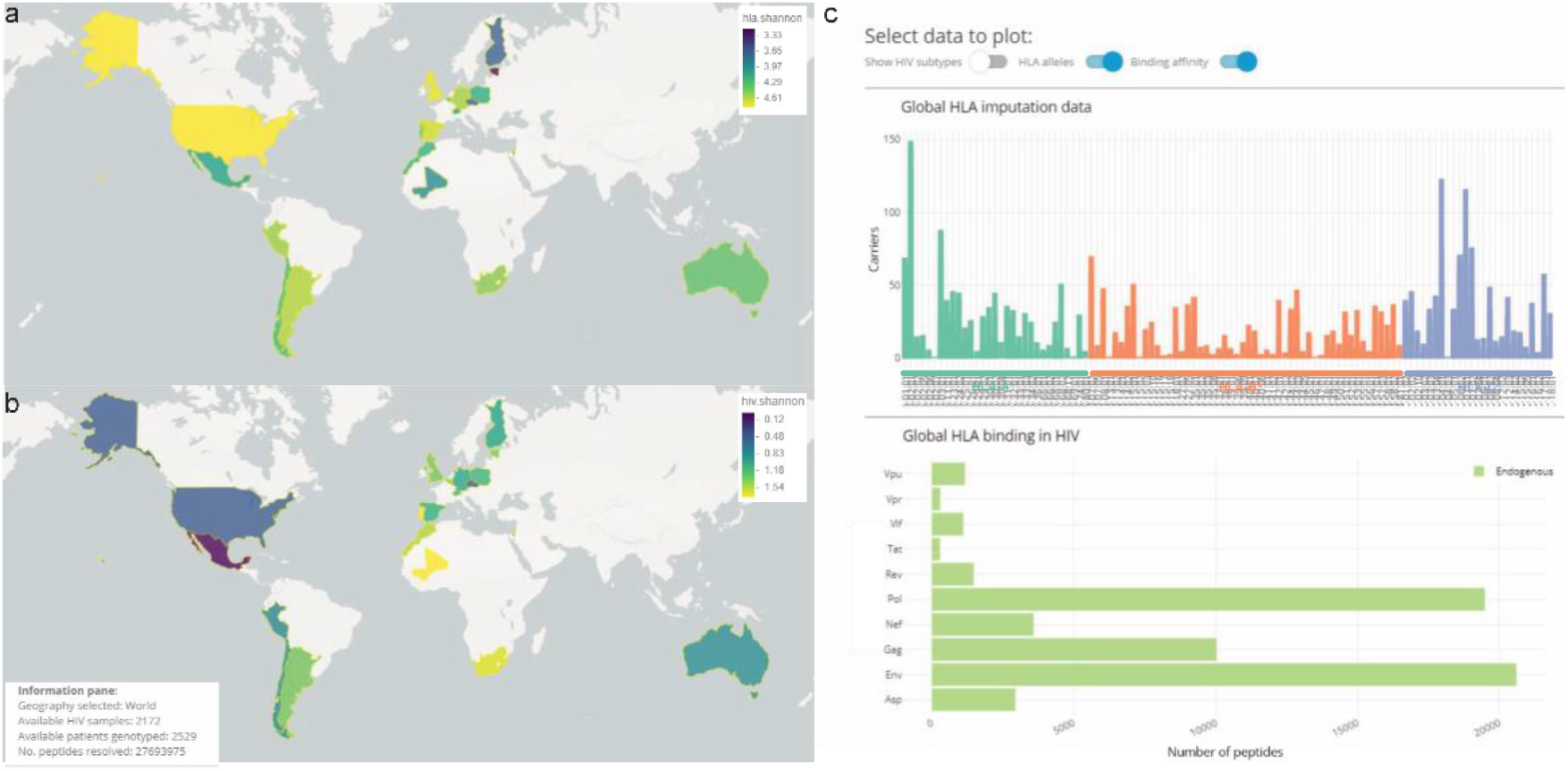
Web application demonstrating a subset of available data visualizations. Panel (a) and (b) illustrates the global diversity of imputed HLA alleles in Shannon and Simpson indices, respectively. Darker (navy) colors indicate higher diversity. Panel (c) showcases the interactive component to further examine specific HLA allele, an HIV subtype and/or peptide frequencies for the selected cohort with options to explore individual countries in detail.

### Higher HLA-A diversity is associated with a lower mean viral load per country

The diversity of HLA class I alleles measured in terms of the inverse Simpson index was calculated using the HLA allele frequencies per locus and country for comparison. A negative univariate correlation (R = −0.65 p = 0.0018) between HLA-A diversity and mean HIV log_10_(viral load) per country was found. This indicates that countries in our cohort with a high diversity of HLA-A alleles would show lower levels of HIV viral load in the population represented in terms of mean log_10_(VL). No significant correlation was observed for HLA-B and HLA-C respectively (Figure 2).

**Figure 2.**
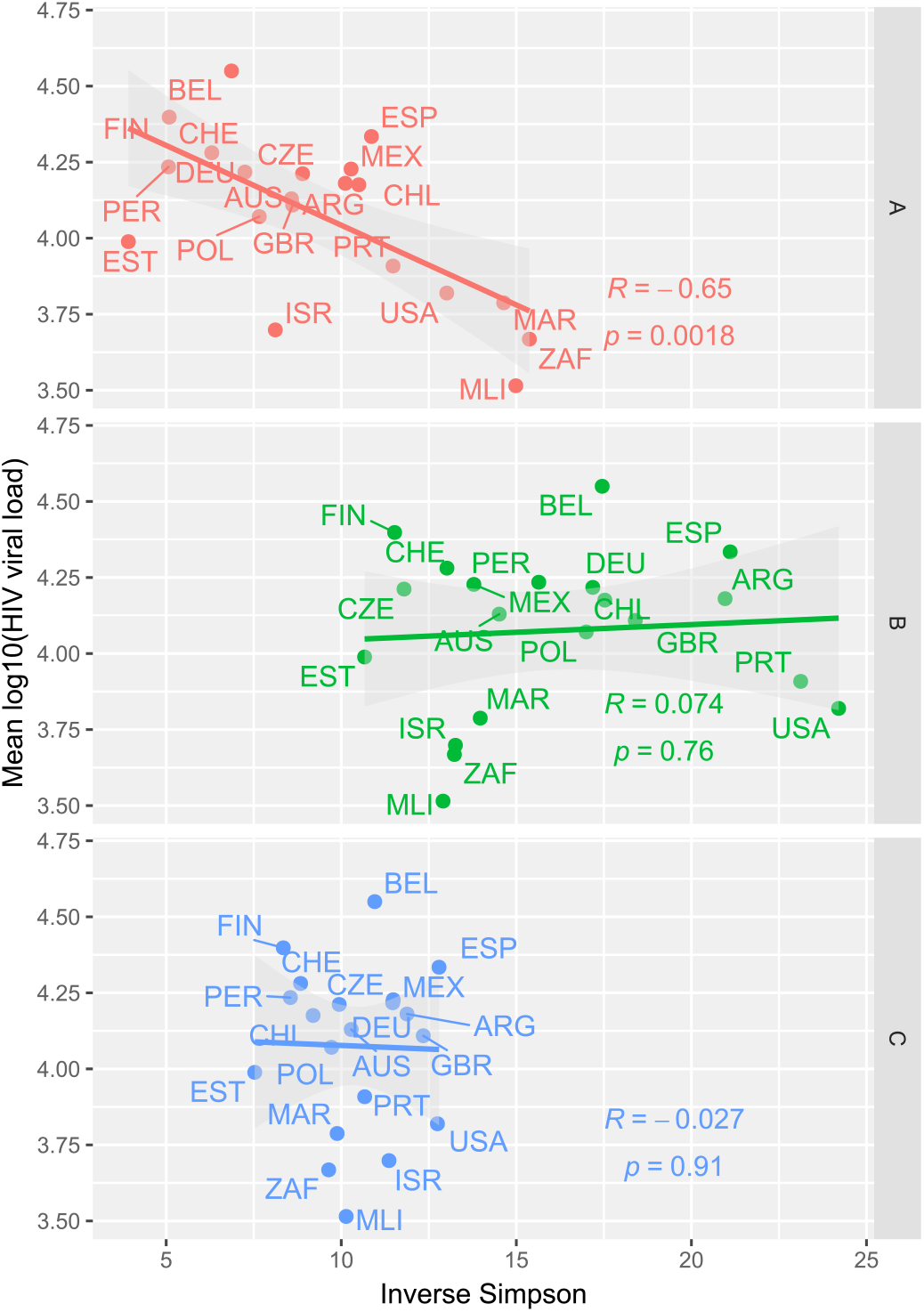
Correlations between HIV viral load and HLA diversity per country for three HLA loci. Mean log_10_(HIV-VL) was depicted (y-axis) as dot plots against HLA diversity measured in terms of Simpson index (x-axis) for three HLA class I loci (HLA-A, -B, -C) in 21 countries. The noted R corresponds to a Pearson correlation with its corresponding p-value.

### Consensus clustering of HIV-derived immunopeptidomes improves statistical power compared to random immunopeptidomes

Consensus trees were generated based on a half-million random peptides from Uniprot and the predicted HIV immunopeptidome under the same methodology. The correlation between both trees was 0.985 indicating high similarity. Minor differences were found in allele distances (Figure S1) with only a few major structural changes such as the displacement of B*35:20 from a predominantly B*15 node (unspecific immunopeptidome) to one dominated by B*35 alleles (HIV immunopeptidome). Another two examples are HLA-B*15:13 and B*15:58 which moved from a node of predominantly HLA-C alleles (unspecific immunopeptidome) to a node of HLA-B alleles (HIV immunopeptidome). These small differences, when combined, were translated into an increase in statistical power (Figure S2). Overall, when assessed in a multivariate model including all HLA class I alleles as covariates the combined percentage of explained variance accounted for 11.44% of the VL after adjustment for sex, self-reported race, and country. Hence, our clusters of HLA function accounted for HIV-VL variance to a similar degree as reported by Bartha et al., 2017 [3] for genotypic evidence in a comparatively pure European cohort. This highlights that accounting for HLA function is both sufficient and better to explain variability in VL in heterogeneous populations.

### HLA class I functional nodes associations with HIV-VL

Associations between the HLA functional nodes and the measurements of HIV-VL taken at study entry were tested using linear regression. Four nodes with a consistent effect size were associated with HIV-VL (Figure 3). These nodes were composed of 30 HLA class I alleles of which 11 were observed in participants with measurable VL at study entry. Two functional nodes were associated with a lower VL, one group composed of HLA-B*57:01, B*58:01, B*57:02, and B*57:03 (β −0.25, q-value 7.02E-06) and the second group composed of a pair of HLA-C*08 alleles, HLA-C*08:04 and C*08:01 (β −0.29, q-value 0.042). In contrast, two nodes showed an association with higher VL, one cluster composed of six HLA-B*44 alleles, B*44:05, B*44:08, B*44:04, B*44:03, B*44:02, B*44:27 (β 0.15, q-value 0.003) and a mixed group composed of 16 alleles: B*35:20, B*35:16, B*35:10, B*35:43, B*35:08, B*35:19, B*35:41, B*35:01, B*35:17, B*35:05, B*44:06, B*56:03, B*53:01, B*15:08 and B*15:11 (β 0.13, q-value 0.048). From these alleles, only two (HLA-B*57:01 and B*57:03) were associated with HIV-VL when tested at the individual allele level (Figure 3).

**Figure 3.**
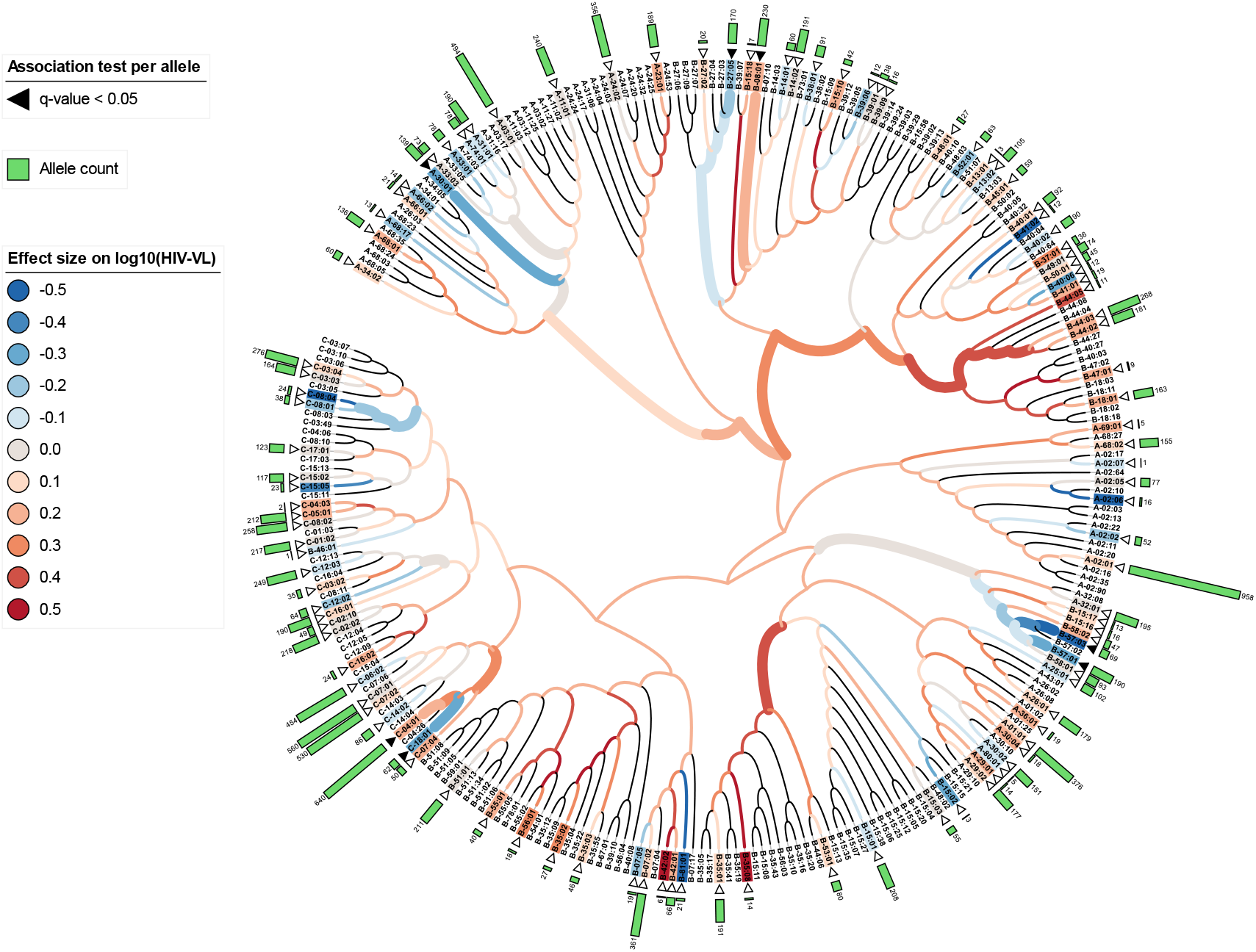
Dendrogram of 268 HLA class I alleles based on consensus clustering of predicted binding affinities to HIV peptides. Predicted binding affinities to 173,792 HIV peptides were used to calculate the HLA allele distances used for consensus clustering and represented as a dendrogram through hierarchical clustering. Associations with log_10_(HIV-VL) of each node (HLA functional node) and leaves (HLA alleles) in the dendrogram were tested and adjusted by sex, self-reported race, and country. Associations were defined by an adjusted p-value (Benjamini-Hochberg) < 0.05 and are represented as thick branches for nodes and black triangles for leaves. White triangles indicate HLA alleles detected in our cohort. The effect of the respective associations is color-coded from protective effect (blue) to detrimental (red). On the outer ring, HLA allele counts are depicted as green bars. An interactive version of the tree can be found in the provided web application.

## Discussion

In this study, we implemented a computational approach based on consensus clustering of HLA alleles using predicted immunopeptidomes to explore associations of functional HLA groups in a geographically diverse cohort of PLWH. We defined functional similarity as differences in epitope binding profiles between HLA alleles and performed HLA imputation on patients enrolled in the START study. We combined the host genetic information with viral genomics through the prediction of immunopeptidomes. Viral sequences derived from a subset of participants were processed into peptides to predict binding affinities to 268 HLA class I alleles which allowed us to generate distance matrices of HLA alleles. Consensus clustering allowed us to agglomerate HLA alleles into nodes by their functionality. Four nodes were found to be associated with HIV-VL, implicating alleles that could not be detected when performing independent allele-specific tests alone. These four nodes accounted for a total of 30 HLA alleles of which 11 were observed in our cohort.

The effects differed among the four nodes associated with HIV-VL. One node containing four well-characterized alleles HLA-B*57:01, B*58:01, B*57:02, and B*57:03 showed an association with lower HIV-VL indicating a protective effect. This effect has been previously reported on an individual allele level [28]; we confirmed and observed shared binding affinity profiles that now indicate their common mode of function. While having a similar effect in HIV-VL, these four alleles are represented at different frequencies between populations. For example, HLA-B*57:01 carriers are of European descent compared to B*57:03 which is predominant among those of African descent [2]. The similarity in function was captured by our analysis despite the differences in allele frequencies. A second protective node contained two alleles, HLA-C*08:04 and C*08:01, which have been reported in an admixed population and detected after adjusting for multiple factors such as CD4+ and CD8+ counts [29]. We showed that such association could not only be detected in our study but that functional clustering provided a clear and simple method to do it with greater mechanistic insight. We report a node of six HLA-B*44 alleles, in this case, associated with a higher VL. This finding contradicts a previous study in a Chinese cohort [30] in which a few alleles of the cluster were found to have a protective effect. Given the large effect of viral diversity on VL, this could be a result of regional adaptation that could not be accounted for in our study due to the lack of participants from the same region. While this suggests a weakness in our study due to the underrepresentation of some ethnicities, divergence in our results from studies on homogeneous populations could be a useful tool to detect localized effects. However, data from a cohort with sufficient Chinese representation is needed to confirm the findings.

Defining discrete nodes of HLA alleles is challenging as distances between alleles are based on predicted binding affinities and measured continuously. Therefore, metrics to define a fixed number of clusters failed to suggest a robust threshold for cutting the trees obtained by consensus clustering. For this reason, the final HIV-associated groups of alleles were determined by assessing the trees hierarchically from leaves (individual HLA alleles) to roots (broad HLA nodes), thus optimizing groups based on their association with VL when the combination of alleles in all child nodes supported it. In this way, the methodology implemented provides an advantage to traditional HLA association studies by adaptively increasing the signal of functional groups allowing to uncover associations that could not be found at the individual allele level due to the number of statistical comparisons or sample size [14]. Grouping or clustering of variables to increase statistical power and limit multiple testing is common practice in epidemiological studies. The functional clustering presented here, however, is based on the assumption that the majority of HLA functionality is mediated by peptide binding affinities. This serves to facilitate study design by simplifying decisions on the size and diversity of the cohort while allowing a biological interpretation of the results. For example, it enables the inference of the effect of unobserved HLA alleles on HIV-VL given their common function to those observed. These are notable challenges across clinical studies attempting to measure the effect on the immune response. Altogether, the functional clustering approach presented is neither specific to HIV nor viral load and may be transferred to study HLAs and outcomes in other host-pathogen interactions.

Similar computational approaches were proposed to explore HLA class I molecules and their role in HIV-1 infection: These studies mainly focused on genome-to-genome approaches [6], techniques to find new epitopes in European participants using a peptidome approach [7] or to explore the interactions of HLA class I molecules to other ligands [31]. In contrast, we focused on the host factors, the HLA class I alleles, accounting for the high diversity of the cohort and inclusion of a larger number of viral sequences to predict immunopeptidomes and expand on genome-to-genome analyses previously performed in our same cohort [19]. While implementations of HLA clustering in the context of HIV-1 have been developed based on a Bayesian framework [13] these were limited by the diversity of their data and assumptions of the clustering parameters. Extra considerations must be taken when clustering predicted immunopeptidomes to avoid bias introduced by predominantly low binding affinity predictions, which can affect the reliability of the computed distances between HLA alleles. To solve this, we used consensus clustering by computing different subsets of immunopeptidomes and avoiding bias from non-binders when calculating distances between HLA class I alleles under multiple linkage functions. The ensemble of implemented techniques avoids bias due to high dimensionality and has shown success when applied to clustering of biological data [22].

We propose diverse methodological improvements in the analysis of host-viral genetics. From the host genetics perspective, we showed that imputed HLA class I alleles can be used to calculate HLA diversity without requiring full HLA sequences. From the viral genetics perspective, we incorporated viral sequences using a fast k-mer approach to avoid generating a consensus sequence, especially for pathogens of high genetic variability and to take into account the intra-host viral diversity [32]. When combining host and viral information through predicted binding affinities, we showed that the differences between clustering based on specific HIV and random immunopeptidomes facilitate a slightly higher resolution of the trees. We suggest that the unspecific HLA clusters from the latter approach could be used for other infectious phenotypes. This is likely due to the diversity of viral genomes present in our dataset, and this may not be the case for smaller, more homogeneous cohorts. However, it suggests that our immunopeptidomes could work as a proxy for other similarly diverse studies that do not have access to paired viral genomes. Alternatively, new immunopeptidomes may be proposed based entirely on synthetic data at the risk of increased type-II error.

While we demonstrate the utility of the implemented methodology on HIV, there is also a clear road to extend it to the analysis of other pathogens eliciting class-I immune responses. Using this common functional framework, there is scope to analyze the variation of HLA structure in populations and within the context of multiple pathogens. Another possible application could be as a screening tool for the most effective peptides either for targeting populations carrying specific allelic distributions or maximizing coverage of vaccines across geographies. Finally, we propose that the focused consideration of HLA function in terms of peptide binding affinities provides a promising approach to inform modern vaccine design. This would also be relevant in the advent of mRNA vaccines coming to fruition after decades of research [33] as they take advantage of proper peptide selection. As an example, the need for a new class of tools that account for both variable immunogenic coverage and clinically relevant mutations has been highlighted during the recent SARS-CoV-2 pandemic [34]. To facilitate open research, the results of this work are made available for common use via a web application (https://persimune-health-informatics.shinyapps.io/PAW2022ZuccoHLA_HIV_INSIGHT/)

## Data Availability

All data produced in the present study are available upon reasonable request to the International Network for Strategic Initiatives in Global HIV Trials (INSIGHT)

https://persimune-health-informatics.shinyapps.io/PAW2022Zucco__HLA_HIV_INSIGHT/

## Acknowledgements

We would like to thank all participants in the START trial and all trial investigators. See N Engl J Med 2015; 373:795–807[16] for the complete list of START investigators.

## Author contributions

A.G.Z., J.D.L. and C.R.M conceived the study. A.G.Z., M.B, M.G. and C.E prepared the data. A.G.Z performed the statistical and computational analyses. A.G.Z. and C.R.M drafted the manuscript. All authors contributed to data interpretation, critically revised the manuscript, and approved the final version.

