## Supplementary figures for "Associations of functional HLA class I groups with HIV viral load in a heterogeneous cohort"

Adrian Gabriel Zucco, MSc, PhD student

mail:

Rigshospitalet, Copenhagen University Hospital

Centre of Excellence for Health, Immunity and Infections (CHIP) & PERSIMUNE

Blegdamsvej 9, DK-2100 Copenhagen Ø, Denmark

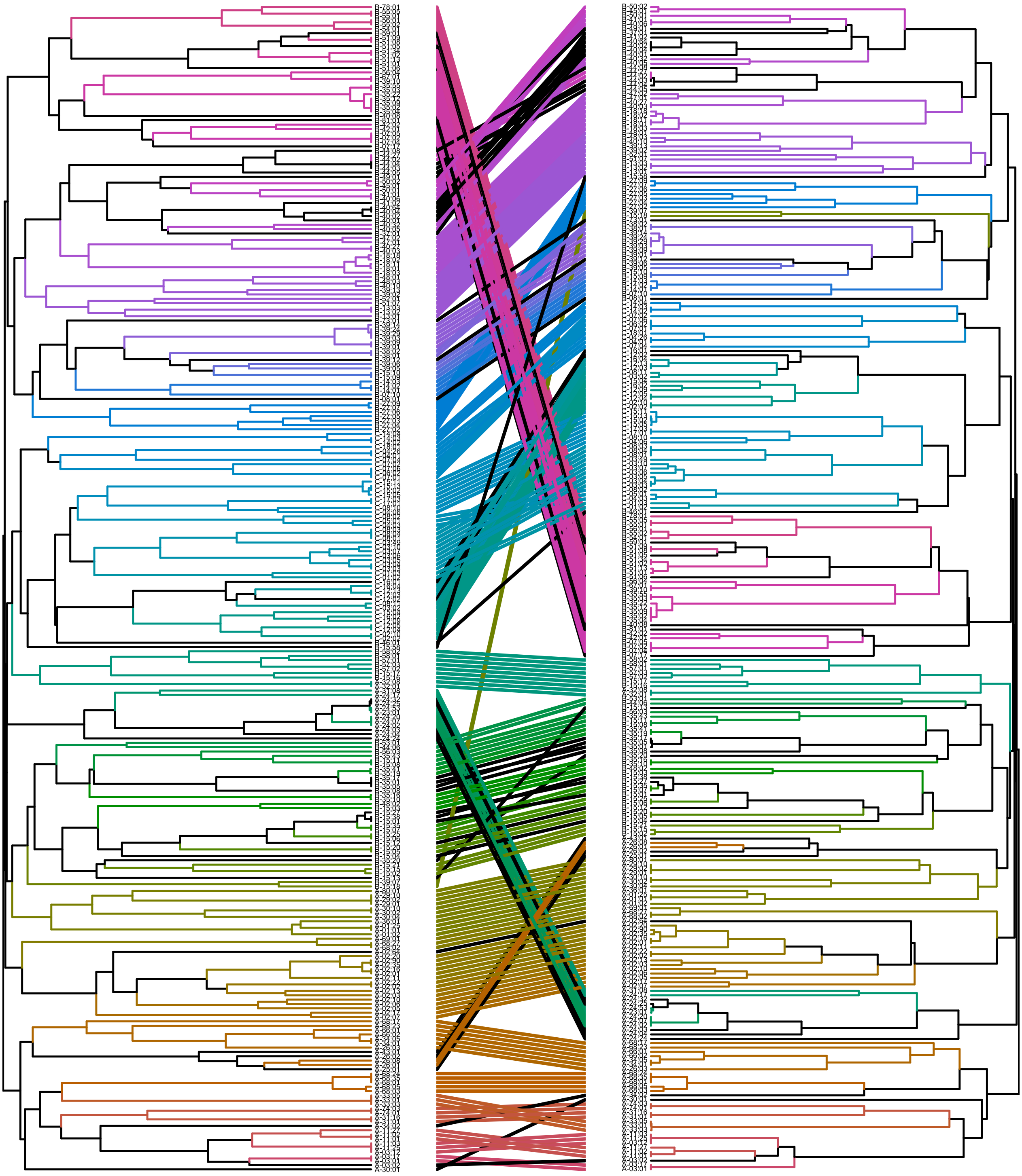

**Figure S1. Tanglegram of consensus clustering from predicted immunopeptidomes based on random versus HIV-specific peptides.**

Two different peptide sets were used for consensus clustering based on predicted immunopeptidomes to 268 HLA class I alleles. On the left, a dendrogram generated from $5x10^5$ random peptides is compared to a dendrogram generated from 173,792 HIV peptides. Black branches and lines connecting both dendrograms indicate differences in clustering among both dendrograms.

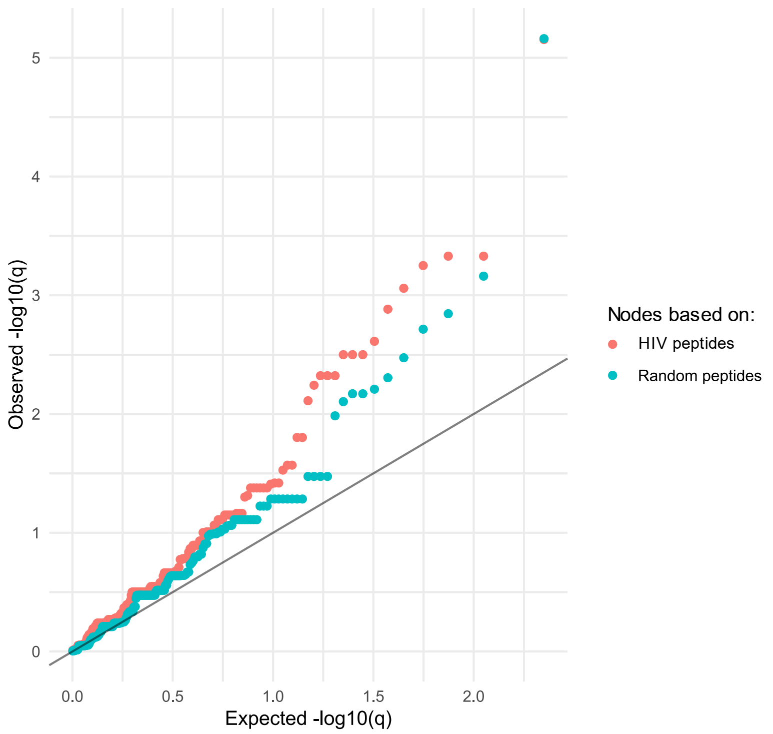


**Figure S2. Q-Q plot of observed versus theoretical q-values from associations to HIV-VL from HLA functional nodes**

HLA functional nodes were generated by consensus clustering of predicted HIV-specific immune peptidomes (red) and unspecific predicted immunopeptidomes from random peptides (blue).
